# A biological variation-based approach to the day-to-day changes of D-dimer, Fibrinogen, and Ferritin levels that are crucial in the clinical course of COVID-19 in healthy smokers and non-smokers

**DOI:** 10.1101/2021.10.24.21265444

**Authors:** Aydın Balci, Muhammed Emin Düz, Elif Menekşe

## Abstract

**Objective:** D-dimer, ferritin, and fibrinogen parameters in COVID-19 patients are essential, particularly in inpatients and intensive care unit patients. It is vital to know the changes that occur due to the biological structure of the person than the disease effect in these tests in order to manage the fatal disease better.

**Method:** Blood samples were taken on the first, third, and fifth days from 30 healthy volunteers, 15 of whom were smokers, 15 were non-smokers, and D-dimer, ferritin, and fibrinogen tests were studied with repeated measurements. After the data processed for normality and homogeneity and removing extreme values, CV_A_, CV_I_, CV_G_, CV_T_, RCV, II, I%, B%, TE% values were calculated via a full nested ANOVA design, according to Callum G, Fraser, and EFLM.

**Results:** CV_I_ and CV_G_ values of D-dimer were calculated as 49.07% and 40.69% for all individuals, 49.26% and 27.71% for smokers, 48.80% and 51.67% for non-smokers, respectively. In terms of fibrinogen, the same analyzes for all individuals were calculated as 11.18% and 10.62%, 3.25% and 20.17% for smokers, 9.11% and 6.79% for non-smokers, respectively. The same analyzes for ferritin were calculated as 23.74% and 63.31% for all individuals, 34.98% and 35.24% for smokers, 30.53% and 74.87% for non-smokers, respectively.

**Conclusion:** Changes in D-dimer measurements every other day in healthy individuals can be observed depending on the biological characteristics of the individuals, and the population-based reference interval may be insufficient for clinical evaluation. Each individual should be evaluated within himself/herself. When evaluating the results of ferritin and fibrinogen in non-smoking individuals, it should be taken into account that significant differences may occur between individuals. Besides, it should be kept in mind that there may be significant changes due to biological variation in the repeated measurements of ferritin every other day.

## INTRODUCTION

The COVID-19 pandemic has opened deep wounds for human history, and its impact continues. Vaccination studies have not reached the desired levels, except for a few countries, and there is no cure for the disease [1]. The number of admissions to hospitals, the rate of inpatients, and the number of admissions to the intensive care unit have not yet fallen to the desired level [2]. For this reason, healthcare providers have to provide rapid diagnosis and treatment and make quick decisions. It is also vital to determine in advance which patient the COVID-19 infection will be mild and which will be severe to protect human life against the deadly virus. In terms of patients, biochemical parameters that can be of great benefit were determined when outpatient treatment, hospitalization, discharge, and admission to the intensive care unit were considered clinically [3-5]. Especially in the follow-up of inpatients and intensive care unit patients, D-dimer, fibrinogen, and ferritin have come to the fore, and they are influential on clinical decisions [6,7].

Although the follow-up of biochemical parameters helps make sense of the clinical course of patients with other examinations and clinical findings, there are other variables that should be taken into account when evaluating patient results. At this point, biochemically essential changes occur within and between individuals due to the nature of the organism and develop independently of diseases. Changes within the population-based reference range, mainly used in medical laboratories, may have clinical significance and are often ignored. Intraindividual variations may be related to natural variation [expressed as coefficient of variation (CV)] consisting of variations in preanalytical (CV_P_), analytical (CV_A_), and within-subject biological variation (CV_I_) [8]. Between-subject biological variation (CV_G_), described as the changes between the setpoints of different individuals, is another component of biological variation (BV) like CV_I_ [8]. The index of individuality (II) calculated in the form of CV_I_/CV_G_ is helpful at the point of whether the use of population-based reference intervals can be beneficial or not [9]. The reference change value (RCV) obtained from the BV data provides information on whether the differences in repeated measurements of the same analyte are clinically significant [10]. Previously the biological variation data compiled and collected by Dr. Carmen Ricos and colleagues on the Westgard website started to be published systematically by the European Federation of Clinical Chemistry and Laboratory Medicine (EFLM) as of May 2019 [11,12]. Besides, at the EFLM 1st Strategic Conference, BV studies were updated and re-standardized by addressing the analysis of BV data, the characteristics of the individuals who will form the sample, the differences in the working groups, and the uncertainties in the calculations [13]. Above all, EFLM has continued to work to accomplish the challenges on these problems and has strived to broaden the reliable BV data by courtesy of the Working Group in BV (WG-BV) [14]. Also, analytical performance specifications (APS) for imprecision (I%), bias (B%), and total error (TE%) of several analytes derived from BV data were published on the EFLM website [12].

It has been reported that for biological variation analysis, samples should be taken from healthy individuals within a period of at least ten weeks, weekly [15]. However, it is essential that we must use all the resources we have in combating the deadly and incurable COVID-19 pandemic. Between-individual and intra-individual changes in follow-up parameters directly affect clinical decisions about patients. If CVI, CVG, and RCV can affect these decisions on a daily basis, it may be important to evaluate the results from this perspective in terms of benefit to patients. This is our primary motivation that drives our work to realize.

## MATERIALS and METHODS

### Subjects

Thirty healthy individuals as fifteen smokers and fifteen non-smokers, volunteered for the study. Volunteers were selected from the laboratory work team. A detailed explanation about the study was made to the volunteers. Informed consent was obtained from individuals. Amasya University ethics committee approval was taken for the study. The reason to form the groups according to smoking is because of the prothrombotic, inflammatory, and endothelial damage effects of smoking. Since these effects can impact our research parameters and smoking is an essential factor in COVID-19 patients, we carried out our group planning in this direction. Unlike the known biological variation studies, samples were taken from the volunteers for a total of three days, on the first, third, and fifth days, as the parameters that affect the clinical decisions taken in terms of daily changes in COVID-19 were evaluated. Those with a history of deep vein thrombosis and pulmonary embolism, and those who received antiaggregant or anticoagulant therapy were excluded from the study, as their D-dimer and fibrinogen results would be affected, and those with chronic inflammatory conditions, anemic patients, and those receiving iron therapy excluded because their ferritin levels could be affected. Those who had COVID-19, pregnant women, and regular medication were also excluded from the study.

### Study Design

Fasting blood was collected from the volunteers every other day, on the first, third, and fifth days, in a yellow-capped tube with no gel additive and a blue-capped citrate tube. Blood samples were drawn between 8 a.m - 10 a.m with an 8-hour night fasting by the same trained phlebotomist. Twenty minutes after blood was taken, plasma and serum samples were obtained by centrifugation at 1500 G for 15 minutes. Plasma and serum samples were accumulated at -80 ° C until the day of the study. After waiting for the samples to dissolve at room temperature on the analyzing day, centrifugation was performed again at 1500 G for 15 minutes. All tests were duplicated to assess analytical variation. Serum ferritin analyzes were performed on a Siemens Advia Centaur XPT immunoassay autoanalyzer (Siemens Healthineers AG, Erlangen, Germany) with a reference range of 10-322 ng/mL, 2.1-3.0 within run % CV, and 2.7-5.4 between run % CV. Fibrinogen levels were analyzed with Stago Compact Max 3 automatic coagulation analyzer (Diagnostica Stago, Inc.Parsippany, NJ, USA) with the Claus coagulation method, with a reference range of 200-400 mg/dL, 1.60- 1.72 within run % CV, and 3.77-4.73 between run % CV. Plasma d-dimer levels were studied on Erba XL 1000 clinical chemistry analyzer with a turbidimetric method (Erba Corporate Services, London, United Kingdom) of Archem brand reagent (Archem Health Ind. Inc. Başakşehir, Istanbul, TURKEY). The 99th percentile limit is specified as ≥0.5 µg FEU/mL, within run % CV 1.3-3.7, between run % CV 3.3-4.5. All analyzes were performed on a single day with the same lot number of calibrators, controls, and reagents. On the days of the studies, two levels of quality control samples, one normal and one pathological were run for each analyte to verify suitability for analysis. In order to reduce analytical variation, the assays of all volunteers were run under the same calibrator curve.

### Statistics

Between replicates, within individuals, and between individual values were analyzed, and whether the data conformed to the normal distribution was found with the Shapiro-Wilk test. The Dixon-Reed criteria was assessed to detect outliers in mean between-subject values for analytes. Besides, Cochran’s C test was used to exclude outliers from within-subject values, including duplicate measurements. After processing the results for extreme values, the homogeneity of variances was analyzed using the Bartlett test applied to all data, including repeated measures. Subsequently, according to Fraser CG, analytical (SD_A_), intra-individual (SD_I_), inter-individual (SD_G_), and total (SD_T_) standard deviations were calculated using a nested ANOVA design. Then results were converted to coefficients of variance component (CV_A_, CV_I_, CV_G_, and CV_T_, respectively). Calculated SD values were converted to CV values using the formula [CV = (SD / mean) * 100]. Besides, reference change value (RCV) and index of individuality (II) values were calculated. Statistical analyses were made using Excel 2016 (Microsoft Inc., Redmond, Washington, USA) and Minitab 19 (Minitab Ltd., Coventry CV3 2TE, UK).

The following calculation was used to calculate the RCV for the parameters:

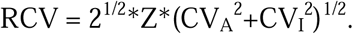

II values were obtained using the following calculation:

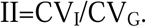

Finally, analytical performance specifications (APS) for the targeted uncertainty (% I), bias (% B), and total error (% TE) are specified using the formulas shown below:

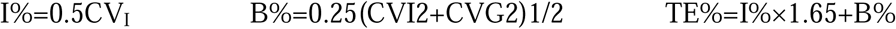

## RESULTS

The data of the 30 patients were statistically analyzed. According to normality tests, D-dimer, fibrinogen, and ferritin parameters were found to comply with the normal distribution (p=0.074, p=0.063, p=0.086, respectively). After performing the extreme value analyzes, individuals 8 and 12 from the non-smoking group and individual 15 (number 30 in total) from the smoker group were excluded from the study together with the repeated analysis data for ferritin. According to the repeated normality tests, ferritin was again found to be normally distributed (p=0.067). After testing the homogeneity of variances (p=0.867 for D-dimer, p=0.643 for fibrinogen, p=0.437 for ferritin), all SD values were calculated over a fully nested ANOVA design, then% CV values were converted. Then, RCV, II, B%, I%, and TE% values were calculated using the formulas indicated. All calculations were made for the whole patient group, the smoker group, and the non-smoker group. The distribution of patient data in terms of parameters was graphically demonstrated in figure 1, figure 2, and figure 3. The results of the biological variation analyzes are shown in table 1.

**Table 1.**
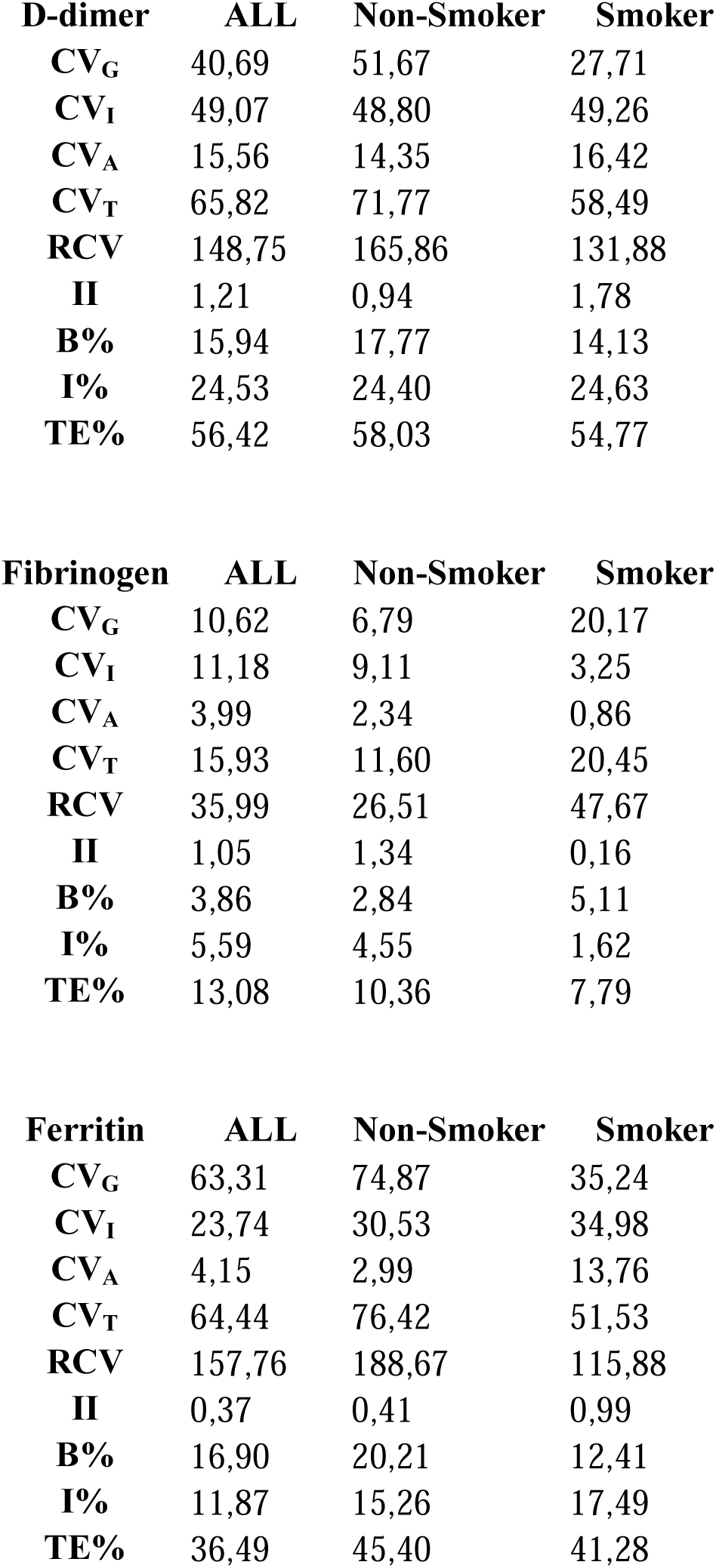
Biological variation analytical performance specification data analyses of D-dimer, fibrinogen, and ferritin measurements.

**Figure 1.**
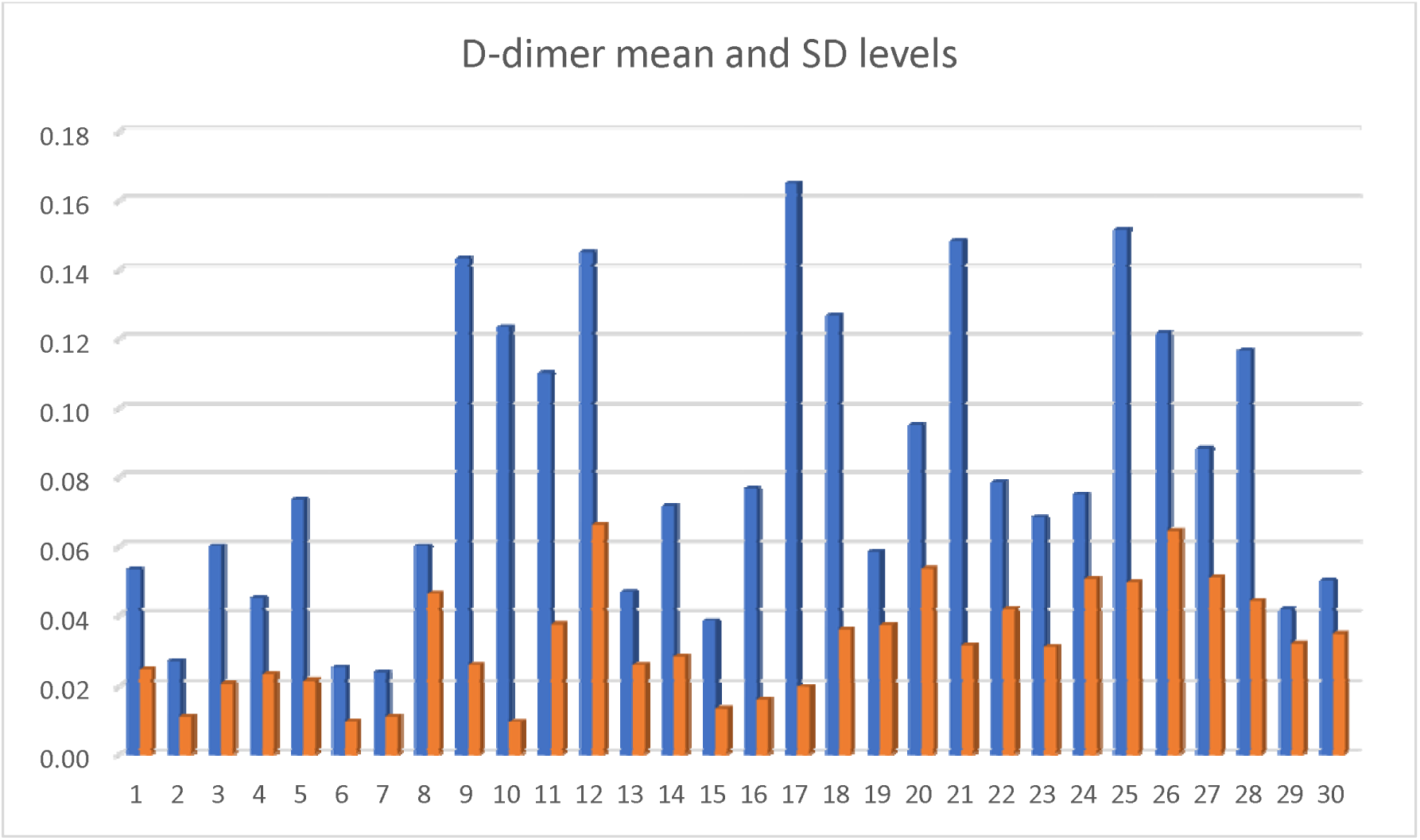
D-dimer mean (µg FEU/mL) and SD levels of numbered individuals. The first 15 people refer to the non-smoker group and the last 15 to the smoker group. Blue bars represent mean, and orange bars represent SD levels.

**Figure 2.**
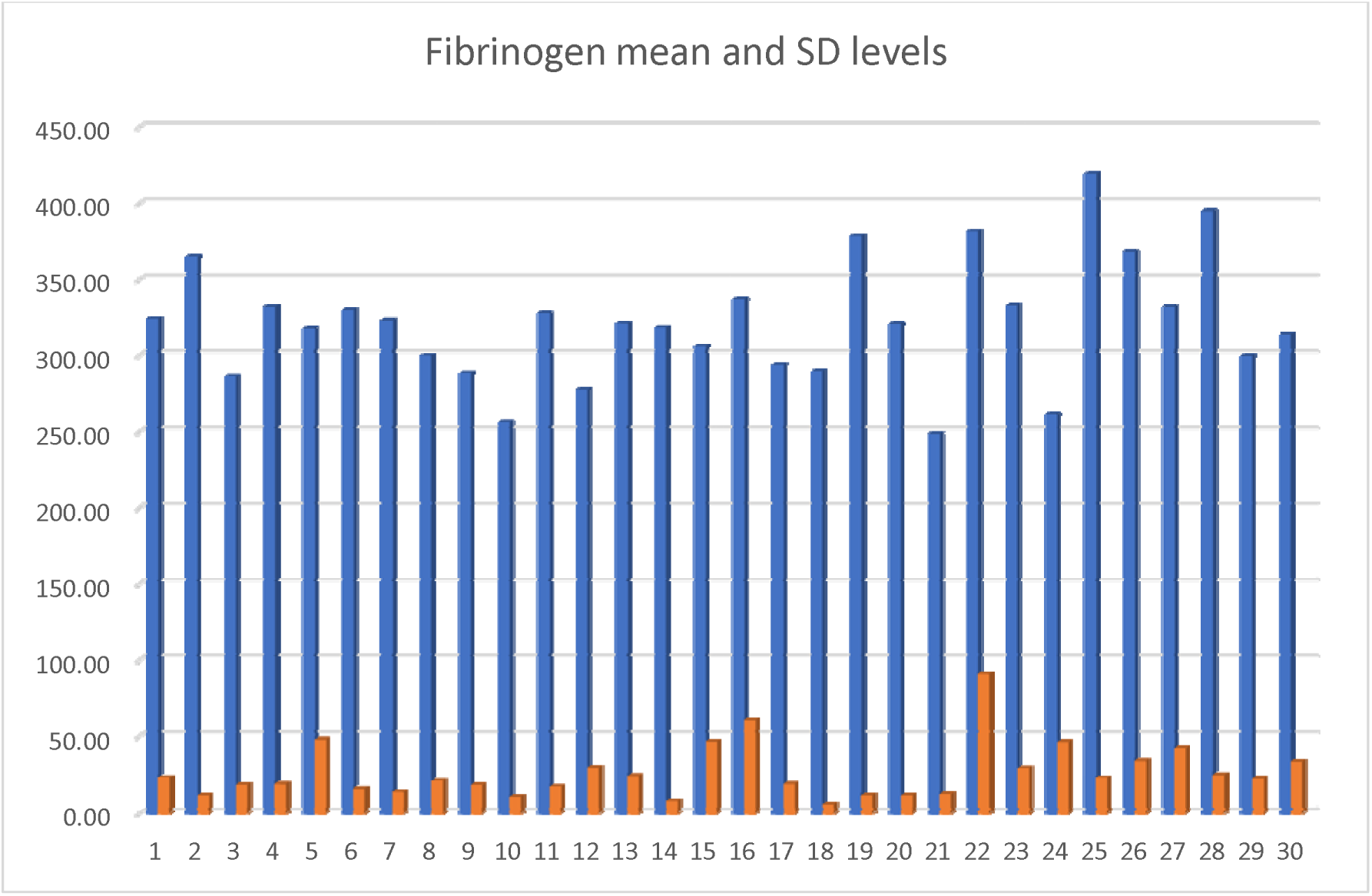
Fibrinogen mean (mg/dL) and SD levels of numbered individuals. The first 15 people refer to the non-smoker group and the last 15 to the smoker group. Blue bars represent mean, and orange bars represent SD levels.

**Figure 3.**
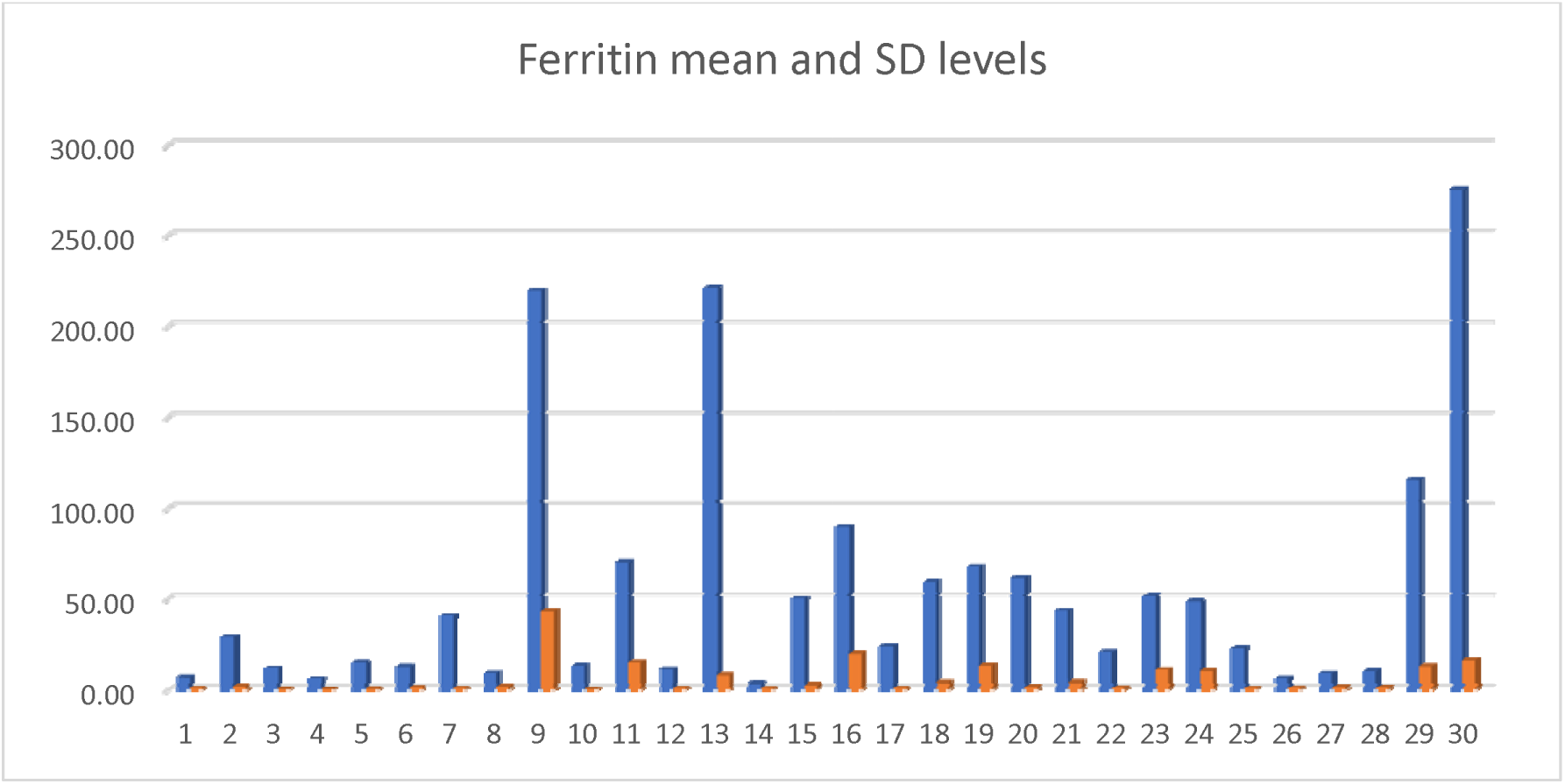
Ferritin mean (ng/mL) and SD levels of numbered individuals before removing the outliners. The first 15 people refer to the non-smoker group and the last 15 to the smoker group. Blue bars represent mean, and orange bars represent SD levels.

## DISCUSSION

Life is a biologically constantly changing process. Laboratory parameters vary between individuals depending on the genetic characteristics of individuals, environmental factors, exposure factors, and many other unknown reasons. Therefore, population-based reference ranges, which are generally used in medical laboratories, may not be sufficient for the clinical evaluation of individuals. Different individual responses to infection may occur, especially in pandemics such as COVID-19 that have not yet been fully enlightened.

The subject is limited in terms of literature regarding BV and APS data of D-dimer. In a study of 19 healthy non-pregnant women, the CV_I_ for D-dimer was 23.3, CV_G_ 26.5, and CV_A_ 31.8 [16]. On the Westgard website, D-dimer BV data is shown as; CV_I_ 23.5, CV_G_ 26.5, I% 11.65, B% 8.82, and TE% 28.04, data probably taken from the same study [11]. Considering our study data, CV_G_ values in terms of D-dimer are compatible in smokers with previous studies. Besides, the CV_A_ was found to be lower than the stated study before [16]. Our study quality was found to be low in terms of APS. It may be because we sampled every other day, as we aim to see short-term changes rather than long-term weekly sampling. However, our CV_A_ values express the quality of our measurements. D-dimer’s high RCV and II values indicate that the test will show high changes in short-range measurements. Also, test individuality is high, and it would not be appropriate to evaluate patients according to population-based reference ranges.

The researchers determined BV values for fibrinogen as CV_I_ 11.9, CV_G_ 17.0, and CV_A_ 2.7 [17]. In one review, CV_I_ values for fibrinogen were reported in the range of 13-67 and CV_G_ values in the range 33-87 [18]. According to Westgard data, CV_I_ was 10.7, CV_G_ 15.8, I% 5.4, B% 4.8, TE% 13.6 for fibrinogen [11]. Our study findings are suitable with previous studies in BV and APS, except for CV_G_ values that we found higher in smokers than literature. When all patients and the non-smoking group were considered, II values were found to be high, while it was found to be low in smokers. We do not understand the low individuality of fibrinogen in smokers. Perhaps, the difference in the amount and type of cigarettes people use may cause different results between individuals, reducing the individuality index.

While the median CV_I_ value of ferritin was given as 12.8 in the EFLM database, in the studies reported on the same page, CV_I_ was stated in the ranges of 9.7-25.8, CV_G_ 16.46-96.53 [12]. BV and APS values for ferritin are reported on the Westgard website as CVI 14.2, CVG 15.0, I% 7.1, B% 5.2, TE% 16.9-21.7 [11]. In a study conducted with a limited number of men, ferritin’s CV_T_ value was determined to be 21.64 [19]. In one study report, BV values for ferritin were calculated as CV_I_ 21.8, CV_G_ 89.8, CV_A_ 3.2, and II 0.24 [20]. While our study data were similar to the literature data in terms of CVG values, our CVI calculations were higher. Similar to our D-dimer results, CVG values were calculated lower in the smoking group, but the II value was also high. In addition, our RCV calculations included high results. According to our calculations, the population-based reference interval may not be sufficient for the clinical evaluation of ferritin values repeated every other day. Also, ferritin measurements in smokers can show individual characteristics. Although our CVA data were at acceptable levels except for the smoking group, our APS calculations could not exhibit an adequate level of quality.

## CONCLUSIONS

Changes in D-dimer measurements every other day in healthy individuals can be observed depending on the biological characteristics of the individuals, and the population-based reference interval may be insufficient for clinical evaluation. Each individual should be evaluated within himself/herself. When evaluating the results of ferritin and fibrinogen in non-smoking individuals, it should be taken into account that significant differences may occur between individuals. Besides, it should be kept in mind that there may be significant changes due to biological variation in the repeated measurements of ferritin every other day.

Our inability to reach sufficient APS values in D-dimer and ferritin parameters maybe because we do not apply a typical biological variation procedure because we are planning every other day for COVID-19, which requires rapid clinical decision-making and can change treatment plans according to daily parameter changes.

## LIMITATIONS

First of all, this study was conducted with healthy volunteers and does not reflect the clinical and biological conditions of COVID-19 patients. Besides, the study is not a typical biological variation analysis planned according to the procedures specified in the EFLM guidelines but is designed to observe the day-to-day change.

## Data Availability

All data produced in the present study are available upon reasonable request to the authors.

https://docs.google.com/spreadsheets/d/1nX71i-EG5bxk9eUDs0UAf1Dp23aqGxo9xtVInsAswXw/edit?usp=sharing

